# Genetic Variant rs1205 is Associated with COVID-19 Outcomes: The Strong Heart Study and Strong Heart Family Study

**DOI:** 10.1101/2023.08.04.23293551

**Authors:** LG Best, E Erdei, K Haack, JW Kent, K Malloy, DE Newman, M O’Leary, R O’Leary, Q Sun, A Navas-Acien, N Franceschini, SA Cole

## Abstract

**Background:** Although COVID-19 infection has been associated with a number of clinical and environmental risk factors, host genetic variation has also been associated with the incidence and morbidity of infection. The *CRP* gene codes for a critical component of the innate immune system and *CRP* variants have been reported associated with infectious disease and vaccination outcomes. We investigated possible associations between COVID-19 outcome and a limited number of candidate gene variants including rs1205.

**Methodology/Principal Findings:** The Strong Heart and Strong Heart Family studies have accumulated detailed genetic, cardiovascular risk and event data in geographically dispersed American Indian communities since 1988. Chi-square tests, logistic regression and generalized linear mixed models (implemented in SOLAR) were used in analysis. Genotypic data and 91 COVID-19 adjudicated deaths or hospitalizations from 2/1/20 through 3/1/23 were identified among 3,780 participants in two subsets. Among 21 candidate variants including genes in the interferon response pathway, *APOE*, *TMPRSS2*, *TLR3*, the HLA complex and the ABO blood group, only rs1205, a 3’ untranslated region variant in the *CRP* gene, showed nominally significant association in T-dominant model analyses (odds ratio 1.859, 95%CI 1.001-3.453, p=0.049) after adjustment for age, sex, center, body mass index, and a history of cardiovascular disease. Within the younger subset, association with the rs1205 T-Dom genotype was stronger, both in the same adjusted logistic model and in the SOLAR analysis also adjusting for other genetic relatedness.

**Conclusion:** A T-dominant genotype of rs1205 in the *CRP* gene is associated with COVID-19 death or hospitalization, even after adjustment for relevant clinical factors and potential participant relatedness. Additional study of other populations and genetic variants of this gene are warranted.

**Author summary:** Considerable inter-individual variability in COVID-19 clinical outcome has been noted from the onset of this pandemic. Some risk factors, such as age, diabetes, obesity, prior cardiovascular disease (CVD) are well established. The possibility of inherited, host genetic risk factors has also been examined and a number validated in very large, population based datasets. The present study leveraged on-going clinical surveillance of CVD and available genetic information in an American Indian population to investigate potential host genetic contributors to COVID-19 morbidity and mortality. Although the number of cases ascertained was relatively small, a novel variant in the C-reactive protein gene was found to be associated with COVID-19 outcome. This protein constitutes a critical component of the innate immune system and *CRP* variants have been reported associated with infectious disease and vaccination outcomes. Improved understanding of the pathophysiology of this infection may allow more targeted therapy.

## Introduction

The ongoing pandemic of Severe Acute Respiratory Syndrome Coronavirus 2 (SARS-CoV-2) infections has taken a devastating toll on American Indian, Alaskan Native (AI/AN) populations, with hospitalization and death rate ratios for COVID-19 (compared with White ethnicity) reported as 2.5 fold; and 2.1 fold, respectively.[1]

A number of demographic and clinical characteristics, such as male gender, socio-economic deprivation, diabetes, cardiovascular disease and obesity, have been identified as risk factors for COVID-19 associated morbidity and mortality.[2] The hypothesis that this increased burden of COVID-19 morbidity and mortality among ethnic minorities is due to the increased prevalence of these co-morbidities in many populations, has been considered; but the proportion of effect attributable to clinical co-morbidities may, be small.[2] In spite of considerable interest in possible host genetic factors that influence the severity of COVID-19 among all patients, there have been few reliably replicating studies in specific racial/ethnic groups.[3]

The expectation that ethnic disparities in COVID-19 outcomes are primarily driven by population specific behavioral or clinical burden of disease may have resulted in less interest in investigating genetic susceptibility among AI/AN communities. This is of importance for the possible identification of therapeutic targets and prevention strategies assisting AI/AN as well as other populations.

The Strong Heart Study (SHS) and allied Strong Heart Family Study (SHFS), comprise the largest, ongoing prospective epidemiological cohort study among American Indians in 12 different Tribal communities located in Arizona, North and South Dakotas, and in Oklahoma. These cohorts have the requisite genetic datasets and ongoing surveillance of medical records for cardiovascular disease (CVD) and risk factors, including COVID-19, to also assess genetic contributions to COVID-19 death and morbidity risks. The present results derive from a case/cohort analysis of SNPs reported associated with COVID mortality or morbidity in other populations.

## Materials and Methods

### Participants and Cohorts

The SHS/SHFS methodology and design has been described previously.[4,5] Diabetes and hypertensive status were defined as previously;[6] and the dichotomous CVD covariate indicates any history of myocardial infarction, coronary artery disease, congestive heart failure, and atherosclerotic stroke and peripheral vasculature disease.[7] While SHS/SHFS medical record surveillance of participants focuses on ascertainment of outcome events and clinical risk factors related to CVD, in early 2020 ascertainment for COVID-19 was begun, with the recognition of a bidirectional relationship between CVD and COVID-19 infection. The physicians conducting medical records reviews identified COVID-19 as either a contributing or definitive factor in death or hospitalization (disease severity). However, medical record review was triggered primarily by CVD related events (for ascertainment criteria, see Lee et al [4]) and a small number of other conditions, but not primarily by infections. Thus, the identified COVID-19 endpoint of hospitalization is probably under-reported, given the surveillance focus on CVD outcomes. However, all deaths associated with COVID-19 during the study period are included in this report.

The study period was 2/1/2020 through 3/1/2023, when medical record reviews ascertaining COVID-19 outcomes were available. We included all participants alive on 2/1/2020 and identified outcome events (either COVID-19 death or hospitalization) during follow-up. There were no specific ICD-9/10 codes used to identify relevant morbid COVID-19 cases, but physician review of the medical record determined COVID-19 case status based on immune measures and clinical diagnoses, with morbid events for review selected on the basis of standard SHS protocol to ascertain incident CVD.[4]

### Rationale for genetic variants selected

A PubMed literature search for candidate variants previously reported to be associated with COVID-19 pathogenesis or clinical outcomes found previously implicated genes in the interferon response pathway,[8,9] *APOE*,[10] *TMPRSS2*,[11] *TLR3*,[9] *ACE*,[12] *FURIN*[13] and the human leukocyte antigen (HLA)[14,15] regions. We also included ABO blood group polymorphisms which have been among the most consistently identified host factors influencing the COVID-19 phenotype.[16-18] The top 17 “hits” of of the Covid-19 Host Genetics Initiative Browser, round 7 identified additional variants of interest.[19]

Thus our literature search developed a total of 73 plausible candidate variants and resulted in 21 polymorphisms that were available from the genetic resources of the SHS, albeit within two subsets of participants that were genotyped in SHS/SHFS substudies. The candidate SNPs and identifying, published references are found in Table 1.

**Table 1.** Variants genotyped, (associated genes) and allele frequency (AF), when the genotype is available.

| Variant (gene) | RISK ALLELE | SHFS AF of risk allele | SHS AF of risk allele | Literature citation |
| --- | --- | --- | --- | --- |
| rs16944971 (FURIN) | C | 0.01 | NA* | [13,20] |
| rs7412 (APOE) | C | 0.02 | NA | [3,21] |
| rs1205 (CRP), alternate allele =T | C | 0.49 | 0.46 | [22-25] |
| rs3091244 (CRP) | G | 0.31 | NA | [26-28] |
| rs3093068 (CRP) | G | NA | 0.01 | [29,30] |
| rs1800947 (CRP) | C | NA | 0.02 | [23,25,29,31] |
| rs201253322 (IRF7) | C | NA | <0.001 | [3,9] |
| rs116302758 (DPP4) | T | NA | 0.02 | [11] |
| rs56179129 (DPP4) | C | NA | 0.002 | [11] |
| rs12329760 (TMPRSS2) | C | NA | 0.20 | [11,13,32] |
| rs150892504 (ERAP2) | C | NA | 0.001 | [33,34] |
| rs1800795 (IL-6) | G | NA | 0.08 | [23,35,36] |
| rs1799752 (ACE) | Insertion | NA | 0.26 | [3,13] |
| rs8176719 (ABO, O allele) | Insertion | NA | 0.76 | [16,37] |
| rs8176746 (ABO, B allele) | G | 0.02 | 0.02 | [38,39] |
| rs10735079 (OAS3) | A | NA | 0.13 | [40,41] |
| rs1405655 (NR1H2) | G | NA | 0.45 | [42] |
| rs1886814 (FOXP4-AS1) | A | NA | 0.41 | [40] |
| rs2109069 (DPP9) | G | NA | 0.22 | [41,42] |
| rs9380142 (HLA-G) | A | NA | 0.32 | [40,41] |
| rs111837807 (CCHCR1) | A | NA | 0.20 | [42,43] |
| rs2071351 (HLA-DPA1) | A | NA | 0.08 | [40] |
| rs529565 (ABO, intron variant) | A | NA | 0.23 | [40] |
| rs10774671 (OAS1) | A | NA | 0.13 | [40] |
| rs61667602 (LINC02210-CRHR1) | A | NA | 0.06 | [42] |

\* NA, Not available in this cohort

Although genotypes were derived from extensive microarray sources, no attempt was made to screen or identify variants of interest on the basis of genome-wide analysis of SHS microarray data.

The only exception to the qualification of previously identified association was the choice of *CRP* polymorphisms as explained below. This was felt to be justified due to the central role of CRP in the innate immune system and dramatic elevation during COVID-19 and other viral infections.[43]

C-reactive protein (CRP) levels are typically thought of as a biomarker for inflammatory response and not as a pathogenetic factor, although inherited *CRP* variants have been associated with, and thus possibly a modifying factor in some infections[22,23] response to vaccination,[26] as well as other conditions such as cancer[44,45] and pre-eclampsia.[46,47] For this reason, we selected 4 available variants related to *CRP* expression for our analysis.

### Laboratory methods and considerations

Clinical, anthropometric and laboratory measures are reported as obtained from the most recent SHS exam prior to the study period. Laboratory methodology for determination of serum creatinine,[48] HgbA1c[49] and high sensitivity C-Reactive Protein (hsCRP)[50] have been reported previously

The SHFS [5] participants were genotyped with the Illumina Human Cardio-Metabo BeadChip microarray (Illumina, San Diego, CA), incorporating approximately 200,000 single nucleotide polymorphisms (SNPs) in loci previously identified as significantly associated with metabolic and CVD traits.[51] These genotypes were generated exclusively from SHFS participants <u>without</u> diabetes mellitus during exams between 1997-2003. Further details related to quality filters and data preparation have been published.[50]

The SHS participants were genotyped using the Illumina Infinium Multi-Ethnic Global-8 vs1. Quality control included filtering variants with call rates <95% and duplicated QC samples. The *CRP* rs1205 variant was genotyped from <u>both</u> the SHFS (N=1,666) and SHS (N=2,114) participants, providing a total of (N=3,780) rs1205 genotypes. This is presented graphically in Figure 1. Except for rs1205 and rs8176746, genotypic data from other variants were available from either the SHFS or the SHS participants (Table 1).

**Figure 1.**
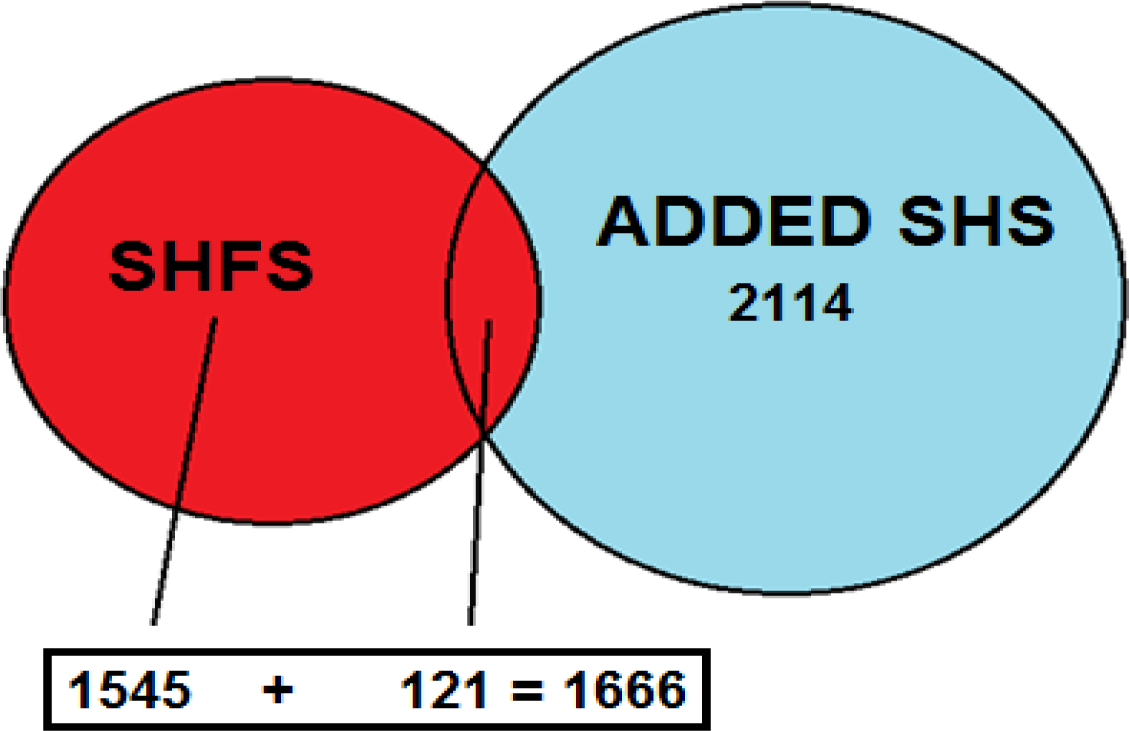
Venn diagram illustrating relationship between analysis cohorts.

Although two different genotyping platforms were used for the SHS and SHFS participants, the genotyping for both was conducted in the same laboratory at Texas Biomedical Research Institute in San Antonio, Texas. All 121 participants genotyped for rs1205 and the ABO variant rs8176746 on both platforms and meeting the criteria for inclusion in the study had concordant genotyping.

### Statistical analysis methods

Descriptive statistics presented counts and percentages for discrete variables and means with standard deviations for continuous variables. Between-group comparison of continuous variables used T-tests and assumed independent samples. Discrete variables utilized Pearson chi-square comparisons between groups.

Logistic regression models were developed for both univariate and multivariate analysis of COVID-19 associated death or hospitalization during the observation period (2/1/20 through 3/29/22). Only a single COVID-event was counted for each participant. To select covariates included in the final multivariate model, BMI and any history of CVD were chosen on the basis of apparent independent association (p<0.05) when all available covariates (sex, body mass index (BMI), hypertension, diabetes mellitus (DM), impaired fasting glucose (IFG) or normal fasting glucose (NFG), history of CVD event, (tobacco (current, ever, never), serum creatinine, and hemoglobin A1c (HgbA1c)) were considered jointly. Age, sex and center were deemed important, standard adjustments and therefore kept in models. The covariate related to diabetes and dysglycemia was not independently associated with COVID-19 when included with BMI (as seen in the first section of Table 5), so was not retained in the final logistic model. The serum hsCRP was not included as a covariate, given we were testing *CRP* genetic variants with known effects on serum levels. Significance was accepted at a p-value of 0.05, since the genetic variants were chosen on the basis of a priori evidence of potential association with COVID-19 as well as recognizing the hypothesis driven nature of the presented analysis.

The SHFS cohort enrolled large families, and therefore we accounted for the relatedness using random effects of pedigree relationships (established with participants at recruitment) in generalized linear mixed models implemented in SOLAR.[52,53] For discrete phenotypes such as affection status, SOLAR employs a classic liability model.[54] Briefly: the unmeasured, presumed multifactorial, basis of liability to disease is modeled as a standard normal distribution (mean=0, SD=1), with affected individuals represented by an upper tail of the distribution that has the same density as the incidence of affection status in the general population. For this study we used the mean weekly incidence of COVID-19 in the US population during the study period (217.72 cases/100,000 population, 0.218%)[55] Heritability of liability is estimated from genetic correlations between relatives[56] and the effects of measured factors (age, sex, etc., as well as measured genotypes) are estimated as deviations of the liability threshold from the population incidence.

### Ethics Statement

All participants in this study have provided written, informed consent to allow genetic and other research related to CVD and its risk factors. Consent was obtained at least once in all cases since Phase I of SHS in 1988, and in most instances has been renewed during each the 6 subsequent phases until the present. Approval has been obtained from the following institutional review boards (IRB) of record: Arizona Area Indian Health Service IRB, MedStar Health Research Institute IRB, Great Plains Area Indian Health Service IRB, Oglala Sioux Tribe Research Review Board, University of Oklahoma Health Sciences IRB, and Oklahoma Area Indian Health Service IRB. In addition to these formal, IRB approvals, all of the participants’ Tribal governments have approved the conduct of the SHS/SHFS studies for these purposes. The close association between CVD (especially stroke) and COVID-19 infection, often in a bidirectional manner, justified IRB approval for this substudy.

## Results

Table 1 shows the genotyped variants selected, their associated genes and allele frequencies in the SHFS or SHS cohorts. The characteristics of both the SHS and SHFS cohorts are summarized in Table 2.

**Table 2.** Demographic and clinical measures stratified by enrolled cohort.

|  | SHFS | SHS | Total | P value |
| --- | --- | --- | --- | --- |
| Participants, N (%) | 1666 (44.1) | 2114 (55.9) | 3780 (100) |  |
| Male Sex, N (%) | 660 (39.6) | 916 (43) | 1576 (42) | .021* |
| Mean Age (SD), years on 2/1/2020 | 52.0 (14.8) | 83.9 (8.4) | 69.8 (19.7) | <.001** |
| Mean Age (SD), years at last clinical exam | 41.7 (14.6) | 62.9 (8.7) | 53.6 (15.7) | <.001 |
| Diabetes status, N, (%) |  |  |  | <.001 |
| Diabetes Mellitus (DM) | 214 (13) | 1001 (49) | 1215 (33) |  |
| Impaired fasting glucose tolerance (IFG) | 347 (21) | 500 (24) | 847 (23) |  |
| Normal fasting glucose tolerance (NFG) | 1080 (65) | 563 (27) | 1643 (44) |  |
| Hypertension, N (%) | 470 (28) | 1110 (53) | 1580 (42) | <.001 |
| Cardiovascular disease***<br>N, (%) | 169 (10) | 870 (41) | 1039 (28) | <.001 |
| Mean BMI (SD), kg/m<sup>2</sup> | 32.3 (7.7) | 30.3 (6.5) | 31.2 (7.1) | <.001 |
| Current smoker N (%) | 644 (39) | 618 (30) | 1310 (35) | 0.657<sup>Δ</sup> |
| Ever smoker, N (%) | 391 (24) | 749 (37) | 1140 (31) | <.001<sup>ΔΔ</sup> |
| Never smoker, N (%) | 619 (37) | 618 (29) | 1237 (34) |  |
| Mean serum creatinine (SD), mg/dL | 0.87 (0.36) | 1.14 (1.22) | 1 (0.90) | <.001 |
| Mean % HgB A1c (SD) | 6.36 (1.6) | 7.10 (2.22) | 6.90 (2.10) | <.001 |
| Mean hsCRP (SD)<sup>****</sup> | 4.12 (3.58) | 4.35 (3.31) | 4.23 (3.45) | .074 |
| COVID-19 any case, N (%) | 48 (2.9) | 43 (2.0) | 91 (2.4) | 0.09 |
| COVID-19 death, N (%) | 21 (1.3) | 26 (1.2) | 47 (1.2) | 0.93 |
| **Participants with either fatal or non-fatal COVID** |  |  |  |  |
|  | **SHFS** | **SHS** | **Total** | **P value** |
| Male sex, N (%) | 18 (37) | 15 (35) | 33 (36) | 0.796 |
| Mean Age (SD), years on 2/1/2020 | 61.9 (14.7) | 78.1 (5.0) | 69.6 (13.8) | <0.001 |
| DM/IFG/NFG, N (%) | 19/5/23<br>(40.4,10.6,48.9) | 17/13/13<br>(39.5,30.2,30.2) | 36/18/36 (40.0, 20.0,40.0) | 0.043 |
| HTN, N (%) | 22 (45.8) | 21 (48.8) | 43 (47.3) | 0.774 |
| CVD, N (%) | 13 (27.1) | 25 (58.1) | 38 (41.8) | 0.003 |
| Mean BMI (SD) | 36.07 (9.10) | 31.04 (6.64) | 33.7 (8.38) | 0.001 |
| Smoking<br>Current/ever/never (%) | 15/15/18<br>(31.3,31.3,37.5) | 14/11/18<br>(32.6,25.6,41.9) | 29/26/36<br>(31.9,28.6,39.6) | 0.828 |
| Serum Creatinine (SD), mg/dL | 0.85 (0.20) | 1.12 (1.86) | 0.98 (1.27) | 0.362 |
| Mean HgbA1c (SD), % | 7.44 (2.14) | 6.65 (1.74) | 7.00 (1.95) | 0.191 |
| Mean hsCRP (SD),<br>mg/L**** | 5.00 (3.78) | 4.48 (3.40) | 4.76 (3.60) | 0.715 |

Values are mean (SD) or N (%). \* Pearson chi square test for count data. \*\*T-test for mean (SD) data \*\*\* Any history of coronary heart disease, stroke, congestive heart failure, or other cardiovascular disease <sup>Δ</sup> smoking "current" vs smoking "never" <sup>ΔΔ</sup> smoking "ever" vs smoking "never" \*\*\*\* hsCRP values >15mg/L excluded

Unadjusted results limited to the primary variant of interest from the SHFS, the SHS cohort or combined SHFS/SHS cohorts, using additive and dominant models based on risk or non-risk alleles are shown in Table 3.

**Table 3.** Primary results of interest, rs1205: univariant associations with either fatal or non-fatal COVID-19.

| Cohort | SNP | Risk allele | Model | Chi-square<br>p value* | OR** | 95% CI | P value |
| --- | --- | --- | --- | --- | --- | --- | --- |
| SHS | rs1205 | C | Add | 0.564 | 0.922 | 0.603 - 1.411 | 0.709 |
| SHS | rs1205 | C | C-DOM | 0.815 | 1.084 | 0.553 - 2.124 | 0.815 |
| SHS | rs1205 | T | T-DOM | 0.368 | 1.452 | 0.642 - 3.285 | 0.370 |
| **SHFS** | **rs1205** | **C** | **Add** | **0.039** | **0.599** | **0.396 - 0.906** | **0.015** |
| SHFS | rs1205 | C | C-DOM | 0.099 | 0.609 | 0.336 - 1.104 | 0.102 |
| **SHFS** | **rs1205** | **T** | **T-DOM** | **0.016** | **2.976** | **1.171 - 7.565** | **0.022** |
| SHS/SHFS | rs1205 | C | Add | 0.066 | 0.743 | 0.553 - 0.998 | 0.049 |
| SHS/SHFS | rs1205 | C | C-DOM | 0.332 | 0.804 | 0.516 - 1.251 | 0.333 |
| **SHS/SHFS** | **rs1205** | **T** | **T-DOM** | **0.020** | **2.038** | **1.105 - 3.758** | **0.023** |

\* Pearson chi-square, asymptotic significance, two-sided
\*\* OR: univariate logistic regression odds ratio

Results for the remaining variants are found in Supplemental Table S1. For variants with low frequency, logistic models were often unstable and not reported. Analyses in the SHFS cohort showed nominally significant associations for the rs1205 T allele dominant (T-Dom) genotype in the combined SHFS/SHS cohort and both additive and T-Dom models in the SHFS cohort).

Table 4 shows the distribution of COVID-19 cases among recruitment centers, highlighting the relatively larger number of cases identified in the Dakota center compared to other centers, and provides the allelic prevalence of rs1205 by center. Among the combined SHFS/SHS cohorts, the prevalence of the rs1205 T allele was 0.525 (95% CI 0.51 - 0.54).

**Table 4.** Combined SHFS/SHS participant and SNP results from adjusted analysis by recruitment center.

| Center* | AZ | DK | OK | p value | analysis |
| --- | --- | --- | --- | --- | --- |
| N | 488 | 1705 | 1587 |  |  |
| Age mean (SD) | 70.65 (19.48) | 68.39 (20.29) | 71.13 (19.05) | <0.001 | 1 |
| Male sex N (%) | 184 (38) | 734 (43) | 658 (41) | 0.104 | 2 |
| BMI mean (SD) | 34.78 (9.88) | 30.25 (6.36) | 31.15 (6.55) | <0.001 | 1 |
| ANY CVD_YN N(%) | 119 (24) | 547 (32) | 373 (23) | <0.001 | 2 |
| SHFS cohort N(%) | 165 (34) | 698 (44) | 803 (47) | <0.001 | 2 |
| Fatal/non-fatal COVID-19 N (% of total) | 11 (12.1) | 62 (68.1) | 18 (19.8) | <0.001 | 2 |
| Risk of outcome % (95% CI) | 2.25% (0.94-3.57) | 3.64% (2.75-4.52) | 1.13% (0.61-1.66) |  | 3 |
| rs1205,C allele count (95% CI) | 0.48 (0.45 - 0.51) | 0.58 (0.56 - 0.59) | 0.48 (0.47 - 0.50) | <0.001 | 4 |
| rs1205 T-Dom (95% CI) | 0.72 (0.68 - 0.76) | 0.81 (0.79 - 0.83) | 0.73 (0.71 - 0.75) | <0.001 | 2 |
| rs1205 T-Dom p value | 0.254 | 0.333 | 0.147 |  | 5 |

\* AZ = Arizona, DK = Dakotas, OK = Oklahoma centers
1= Independent samples, T-test
2= between center Pearson chi-square p value, 3X2 table
3= within center binomial confidence interval on proportion
4= between center Pearson chi-square p value, 3X3 table
5= within center COVID fatal/non-fatal outcome via logistic regression adjusted for age, sex, BMI, ANY\_CVD

Multivariate logistic regression model findings for the primary SNPs of interest are reported in Table 5. Results of the rs1205 T-Dom model is attenuated after adjustment for covariates, but retains nominally significant association (OR 1.859, 95% CI 1.001-3.453, p=0.049) with COVID-19 in in the combined SHS/SHFS cohorts. Within the younger SHFS cohort, the same model showed an odds ratio of 2.857, 95%CI 1.108-7.362, p=0.030.

**Table 5.** Results of multivariate adjusted models evaluating the composite outcome of either fatal or non-fatal COVID-19 within the combined SHFS and SHS cohorts.

| SHFS and SHS COMBINED |  |  |  |  |  |  |
| --- | --- | --- | --- | --- | --- | --- |
|  |  |  |  | 95% CI |  |  |
| Multivariate model including: |  | N | OR | Lower | Upper | P value |
| Age |  | 3780 | 0.999 | 0.987 | 1.012 | 0.923 |
| Sex |  | 3780 | 0.817 | 0.527 | 1.267 | 0.367 |
| AZ center |  | 3780 | Indicator variable |  |  |  |
| DK center |  | 3780 | 2.132 | 1.064 | 4.272 | 0.033 |
| OK center |  | 3780 | 0.649 | 0.296 | 1.423 | 0.280 |
| BMI |  | 3780 | 1.050 | 1.023 | 1.079 | <0.001 |
| ANY CVD (Y/N) |  | 3780 | 1.871 | 1.153 | 3.036 | 0.011 |
| SHFS COHORT |  |  |  |  |  |  |
|  |  |  |  | 95% CI |  |  |
| SNP* | model | N | OR | Lower | Upper | P value |
| rs1205 | C-Add | 1647 | 0.587 | 0.381 | 0.905 | 0.016 |
| rs1205 | T-DOM | 1647 | 2.857 | 1.108 | 7.362 | 0.030 |
| SHS COHORT |  |  |  |  |  |  |
| rs1205 | C-Add | 2042 | 1.057 | 0.683 | 1.634 | 0.804 |
| rs1205 | T-DOM | 2042 | 1.243 | 0.541 | 2.860 | 0.608 |
| SHFS and SHS COMBINED |  |  |  |  |  |  |
| rs1205 | C-Add | 3689 | 0.799 | 0.592 | 1.078 | 0.142 |
| rs1205 | T-DOM | 3689 | 1.859 | 1.001 | 3.453 | 0.049 |

\* Genetic variant included in model adjusted for age, sex, center, BMI, and ANY CVD.

In the SHFS analysis, using a generalized linear threshold model in SOLAR to adjust for the random effect of family relatedness, and adjusting for fixed effects of sex, age, BMI, CVD, and center, either fatal or non-fatal COVID-19 was significantly associated with the fixed effect of the rs1205 T allele-dominant genotype (p=0.0003). The regression beta for this association was negative (= -0.50 standard deviations), indicating a tendency to increase the affected upper tail of the liability distribution (thus confirming rs1205-T Dom genotype as a risk allele).

While those with rs1205 T-Dom genotypes trended toward lower mean hsCRP levels at baseline than others (p=0.087 excluding levels over 15 mg/L to avoid spurious elevations due to intercurrent infections), mean hsCRP levels were not significantly different between COVID-19 cases or controls. (Supplemental Table S2).

## Discussion

We present previously unreported evidence of association between a common human variant (rs1205) and the clinical impact of the SARS-CoV-2 virus causing COVID-19. While our study has a modest number of cases, these results derive from a large cohort of American Indian individuals who are being followed longitudinally using standardized, physician review of medical records[4] The rs1205 variant has been recognized as functionally affecting serum levels of CRP,[25,28,29] in several populations and, in the present SHFS population as well.[50] CRP is an important component of the innate immune system, thus supporting a role of the gene and this variant’s potential effects on the pathophysiology of COVID-19. A genome-wide association meta-analysis has reported an intron variant (rs67579710) associated with COVID-19 hospitalization among 24,741cases and 2,835,201controls.[42] This variant is 4.5Mb from rs1205 [57] and within the thrombospondin3 gene, thus it may play a role in the thrombosis associated with COVID-19, rather than inflammatory pathways.

There are ample theoretical reasons that heritable genetic variation could affect SARS-CoV-2 infection, beginning with the viral requirement for surface receptors to gain entry to the cytoplasm[58] and running through multiple immune response pathways generating the “cytokine storm” which has been prominently noted in the pathogenesis of COVID-19.[59] A number of genetic variants have been associated with morbidity and mortality, including variants in the angiotensin converting enzyme 1 (*ACE1*) gene,[60] the *TMPRSS2* gene,[61] the *IL-6* gene[62] and others extensively reviewed by Ishak et al.[63] The COVID-19 Host Genetics Initiative, a massive ascertainment of nearly 50,000 cases from 19 countries has identified 13 genome-wide loci related to either initial infection, or morbidity.[64]

The rs1205 variant is a C/T single nucleotide polymorphism in the 3’ untranslated region of the C-reactive protein gene (*CRP*), with a prevalence of the C allele ranging from ∼ 0.67 in European populations, to ∼ 0.40 in Asian populations,[65] and 0.51 to 0.54 in the present study. The 4 *CRP* variants were chosen for their role in the innate immune system and availability in the SHS dataset, but no literature was found that examined *CRP* variants related to COVID-19 severity, perhaps because CRP has traditionally been viewed as a biomarker and not as a potential causal factor. There are, however, reports of *CRP* variants associated with the clinical outcomes of other infections[22,23] and the response to vaccination.[26] Additionally, reduced expression of serum CRP is consistently reported associated with the rs1205 T allele, with the beta coefficient for each additional T allele ranging between -0.17 and -0.27;[25,28,29] and estimated at -0.23 in the SHS Dakota center.[50] As noted in Table S2 the mean serum CRP level is lower in the present study, among those with the rs1205 T-Dom genotype compared with those homozygous for the C allele, although the difference is not significant.

Although the prevalent view of COVID-19 pathophysiology implicates a hyperactive immune response (often characterized by increased CRP serum levels as well as coordinated production of other cytokines), it also seems plausible that a relative, baseline insufficiency of an innate immune factor, particularly in the early phase of exposure, could also result in increased morbidity.

*CRP* genotypes correlating with increased baseline CRP levels have been positively associated with risk of pre-eclampsia[46,47,66] and conversely, as in the current study, *CRP* genotypes (including rs1205 T-Dom) correlated with lower CRP levels have been associated with increased infectious burden.[22-24] These divergent effects invite speculation that *CRP* polymorphisms may have experienced balanced selection during evolution.

Our multivariate logistic regression results are in accord with prior studies that showed the contribution of commonly reported clinical factors, such as age,[67] BMI[64] and pre-existing CVD,[68,69] which negatively impact COVID-19 outcomes. Although the mean BMI of the SHFS is somewhat greater than the SHS cohort, it should be noted that the SHFS cohort has a low prevalence of DM (13%) and is younger than the SHFS, due to selection for that substudy of those <u>without</u> diabetes at baseline. Perhaps this allows a clearer genetic (vs an acquired clinical or environmental) association to become apparent.

The small number of cases when stratified by Center suggests cautious interpretation, but is presented since the number of cases from the Dakota Center is considerably higher than from the comparable Oklahoma center. Although the possibility of population stratification confounding our results is present, we believe the fact that all 3 centers show an effect of the T-Dom genotype in the same direction, including the Arizona Center, which is likely to be most distant in their genetic background, suggests genetic background effects are not likely. The statistical significance of the center covariate could be due to various differences in regional, environmental factors, such as household density, weather, and accessibility of medical care.

The lack of significant results in the other SNPs is not surprising, given the number of these variants with low allele frequencies and the small number of COVID-19 cases. Further surveillance of the SHS cohort will likely identify new cases and improve the power for other variants (eg rs111837807 and rs2109069).

The limitations of this analysis are chiefly related to the relatively small number of cases obtained to this point. The morbid case ascertainment is subject to undercounting since the choice of medical records for review is based primarily on CVD outcomes.

This does not affect ascertainment of mortal cases, since all deaths are reviewed and recorded as soon as possible. The possibility of population stratification is noted, although seems unlikely for reasons noted above. P values have not been adjusted for multiple testing.

The study’s strengths include a robust physician assignment of outcomes using medical records and comprehensive analysis of genetic variabilities as well as adjustment for relatedness between individuals.

It is clear that race/ethnicity is a valid and critical risk marker for other underlying conditions affecting the complexity of COVID-19 disease, such as structural racism, discrimination and socioeconomic status,[70] lack of health care access,[71] and exposure to infectious agents related to high risk and service industry occupations.[72] Although one can never be confident that all confounding factors have been adjusted properly, Williamson et al[2] showed that ethnicity among a very large cohort in England was associated with COVID-19 outcomes, even after adjusting for socioeconomic factors. Thus, even though we are committed to improving our understanding of the complex social and behavioral factors influencing COVID-19 disease landscape among Tribal communities we also believe that investigating the potential influence of common genetic variants among the high risk American Indian/Alaska Native populations may provide opportunities for therapeutic or preventive options. Although severely impacted by COVID-19,[73,74] Tribal communities are, at times, excluded from scientific studies due to their smaller proportion in the U.S. population and various socio-political factors.

In conclusion, a statistically significant association was found between the rs1205 T-Dom genotype and risk of COVID-19 death or hospitalization among American Indian participants, employing chi-square tests, logistic regression models adjusting for age, sex, center, BMI and prior history of CVD, and SOLAR software analysis also adjusting for relatedness within the SHFS cohort. The direction of effect suggests a lower level of CRP during early phases of infection may increase the risk of subsequent complications, a novel finding we will continue to investigate further as the pandemic continues and our samples size tragically continues to increase.

Supporting Information:

Supplemental Table 1. Additional univariant associations with either fatal or non-fatal COVID-19.

Supplemental Table 2. Baseline hsCRP measures* in relation to rs1205 T-Dom genotype and COVID-19 case/control status.

## Data Availability

The data underlying the results of this study are owned and controlled by the Tribal entities that approved its collection. This fact is clearly stated in the Tribal resolutions authorizing the research; and it must be recognized that these Tribal communities are an independent, sovereign governments, in control over research activities within their borders. Access to data and materials can be accomplished by application to Mr. Guthrie Ducheneaux, IT director, Missouri Breaks Research Inc, 505 S Willow St, Eagle Butte, SD 57625, 605-964-3419, who will arrange for further consultation with the appropriate Tribal partners. Approximately 2 to 3 months may be required. The authors received special access privileges to the data due to their relationship with the Tribal Governments, however, interested researchers who apply for data access will be able to access the same data as the authors.

## Acknowledgements

We thank the study participants, Indian Health Service facilities, and participating Tribal communities for their extraordinary cooperation and involvement, which has been critical to the success of the Strong Heart Study since 1988.

